# Investigating Climate-Driven Disruption of Malaria Transmission Following the 2022 Sindh, Pakistan Floods: An Interrupted Time Series Study

**DOI:** 10.64898/2026.11.05.26367402

**Authors:** Amna Tariq, Raheel Allana, Sajid Shah, Momin Kazi, Desiree LaBeaud

## Abstract

Malaria remains a major global public health challenge in developing countries, with climate change intensifying its impact, particularly in vulnerable populations. Rising temperatures can expand the geographic range and local abundance of malaria vectors, *Anopheles* mosquitoes, which thrive in warm climates, with 25°C representing the optimal temperature for malaria transmission. In Pakistan, malaria is primarily spread through *P. falciparum* and *P. vivax*. Pakistan is one of the seven countries in the Eastern Mediterranean Region that accounts for 98% of the total malaria burden in the region. Recently, after catastrophic flooding in Pakistan in 2022 and the subsequent increase in malaria incidence, it has become imperative to estimate the climate change-attributed increase in the risk of malaria. In this study, we analyze the time series malaria case incidence data from the province of Sindh, Pakistan, and the climate data to quantify climate-attributed malaria cases in Sindh, Pakistan. We fit a negative binomial generalized linear model (GLM) to monthly case counts with interrupted time series and seasonality terms. The majority of malaria-confirmed infections were Plasmodium Vivax (40-95%) between 2016-2024, followed by Plasmodium falciparum infections. The positivity rate for malaria remained <10% until June 2022, and increased massively after the July 2022 flooding. After accounting for this case momentum, the onset of the 2022 floods was associated with an immediate, independent increase in malaria cases of approximately threefold (IRR 2.985, 95% CI 1.878–4.751, p < 0.001). These findings suggest that post-flooding malaria surveillance and response in flood-affected settings should anticipate not only an acute environmentally-driven surge in cases but also a subsequent period of elevated, self-sustaining transmission that may persist well beyond the resolution of the initiating environmental disruption, in this case the flooding.

## Introduction

Malaria remains a major global public health challenge in developing countries, with climate change intensifying its impact, particularly in vulnerable populations [1]. Malaria has gained significant attention because of its disease burden and its transmission sensitivity to environmental conditions, particularly temperature and rainfall [2]. Variability in climate (rainfall, temperature and humidity) is increasingly recognized as a major determinant of malaria transmission pathways by altering *Anopheles* mosquito reproduction, survival, and the development rate of Plasmodium parasites [3-5]. Literature has shown that rising temperatures can expand the geographic range and local abundance of malaria vectors, *Anopheles* mosquitoes, which thrive in warm climates, with 25°C representing the optimal temperature for malaria transmission [6, 7]. Warmer temperatures accelerate the development of malaria parasites within mosquitoes and shorten the incubation period of the disease in humans, leading to an increased risk of disease transmission.

On the other hand, the relationship between precipitation and malaria transmission is complex. While standing water bodies created after downpour provide a breeding habitat for mosquitoes [8], high-intensity heavy rainfalls may wash or kill the larvae or adult mosquitoes [9]. Studies have reported a non-linear relationship between malaria case incidence and rainfall in Kenya and a positive correlation between rainfall and malaria in Thailand [10, 11].

In Pakistan, malaria is primarily spread through two of the four *Plasmodium* species, namely *P. falciparum* and *P. vivax* [12, 13]. Pakistan is one of the seven countries in the Eastern Mediterranean Region that accounts for 98% of the total malaria burden in the region according to the World Health Organization (WHO) [14, 15]. In Pakistan, around 217 million people are reported to be at moderate risk of malaria, and 63 million are at high risk of the disease [14]. In Pakistan, *An. culicifacies, An. stephensi, An. subpictus, and An. superpictus* are the primary vectors for malaria transmission and have been endemic in the region since the early 1900s [16]. High malaria prevalence in Pakistan is facilitated by the country’s monsoon-fed agricultural landscape, temperatures ranging between 20 °C and 30 °C from September-December and April-May each year, poor vector control and surveillance in the country, and a weak disaster preparedness infrastructure [1, 14].

Sindh Province is one of the five administrative units of Pakistan with a 348km coastline and a population reaching 48 million [17]. Sindh is bordered by the Thar Desert on its eastern side, the Kherthar Mountains on its western side, and the Arabian Sea on its southern side[17]. Sindh is known to have three climatic zones: north, south, and central, and is subdivided into 29 districts. The coastal zone of Sindh is extremely sensitive to climate change [17]. The province is generally located in a dry climate zone with annual precipitation of less than 10 inches and hazards of drought and frequent flooding [17].

Recently, after catastrophic flooding in Pakistan in 2022 [18] and the subsequent increase in malaria incidence, it has become imperative to estimate the climate change-attributed increase in the risk of malaria. In this study, we analyze the time series malaria case incidence data from the province of Sindh, Pakistan, and the climate data to quantify climate-attributed malaria cases in Sindh, Pakistan.

## Methods

### Ethics statement

The study protocol was approved by the Stanford University Institutional Review Board (Protocol ID #31488) and the Ethical Review Board at Agha Khan University.

### Study Site

The study site, Sindh province, is located in the southern region of Pakistan, bordering the Arabian Sea and features diverse landscapes (Fig 1). The fertile plains along the Indus River, with stagnant water bodies, provide optimum breeding grounds for malaria-transmitting mosquitoes. The riverine belt, with its high density of healthcare facilities, experiences a favourable climate for malaria transmission during the wet season (June–August). Conversely, arid regions face significant temperature fluctuations, impacting mosquito breeding patterns. The temperatures frequently rise above 46 °C (115°F) between May and August, and the minimum average temperature of 2 °C (36°F) occurs during December and January. The annual rainfall averages about 18 cm, falling during July and August, which can influence mosquito breeding patterns and potentially affect malaria seasonality [19, 20].

**Fig 1:**
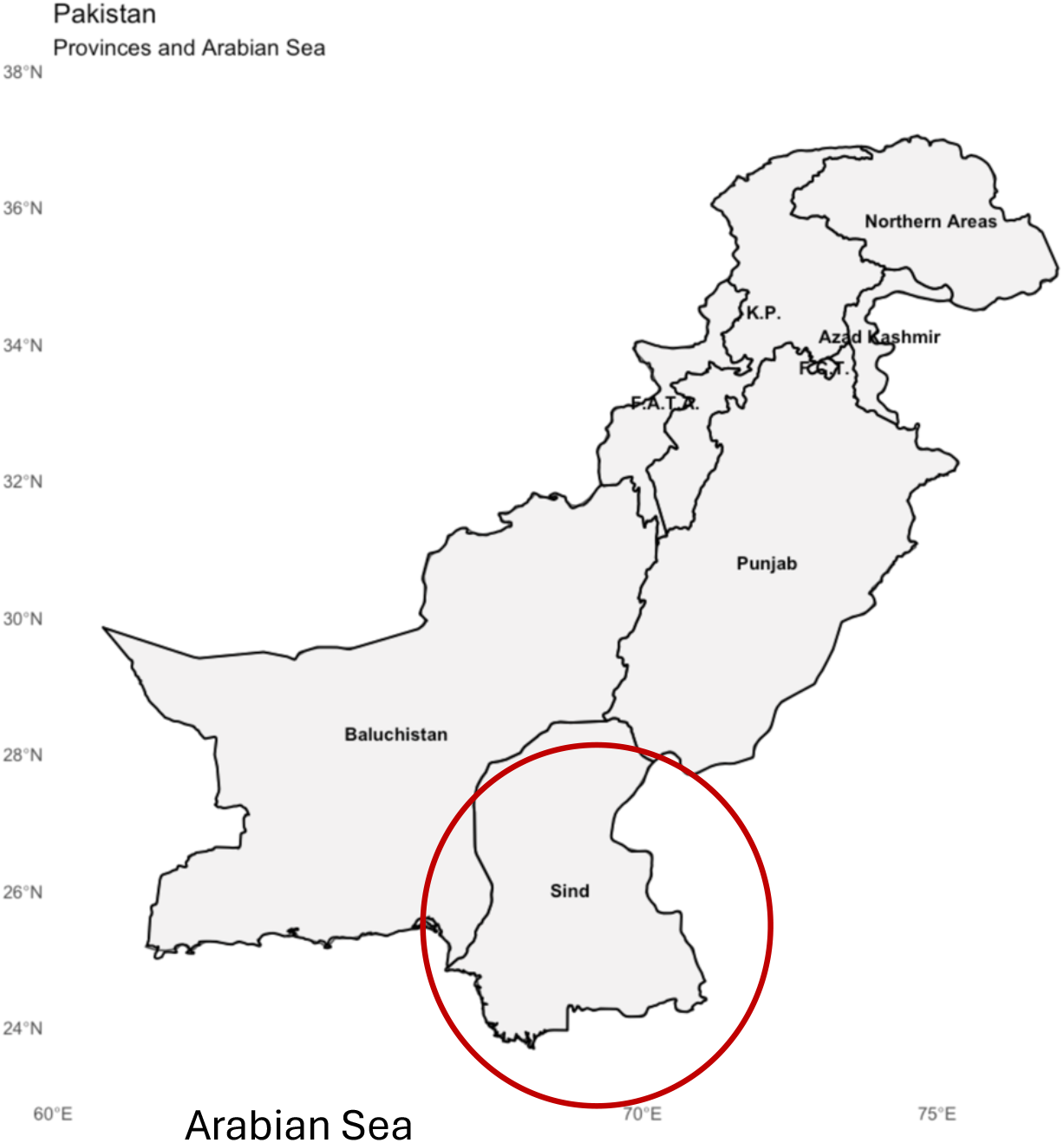
Geographic representation of the study site, province of Sindh.

**Fig 2:**
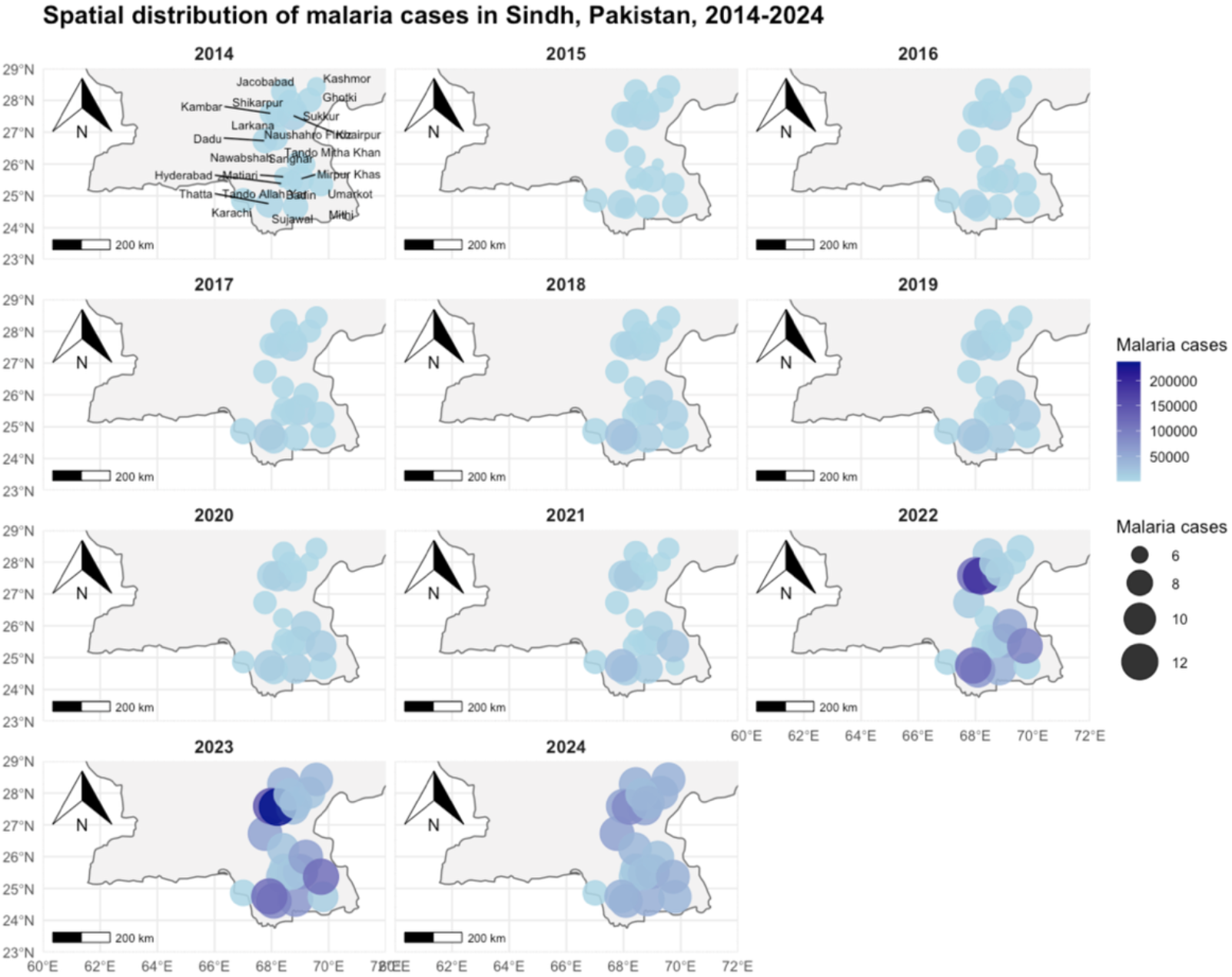
Annual city level malaria incidence from 2014 to 2024.

### Case Data

Monthly longitudinal confirmed malaria cases reported to primary healthcare facilities across 29 districts in Sindh Province, Pakistan, between 2018 and 2025 were analysed. The data were available on a monthly temporal resolution from 2018-2025 with a distinction between malaria caused by Plasmodium Falciparum and Plasmodium Vivax and mixed species.

### Climate Data

The current climate data set is composed of meteorological daily data on mean, maximum, and minimum temperature and precipitation (mm/day), collected and published readily by the “Visual Crossings,” which is an easy-to-use, low-cost source of historical and forecast global weather data [21]. The climate data on temperature and precipitation was temporally aggregated to monthly data to avoid over-parametrization, overfitting, and calculation problems faced with analysing daily climate data.

### Data analysis

Several descriptive summaries (mean, minimum, and maximum) were considered for the two main covariates: temperature and precipitation. The distribution of the mean, minimum, and maximum temperature and precipitation was defined on a daily, weekly and monthly temporal scale, and patterns of temperature and precipitation were identified over time.

### Identification of a structural break

Visual inspection of the case series revealed a marked and sustained increase in monthly case counts beginning in August 2022, coinciding with Pakistan’s 2022 monsoon rains, which caused extensive and prolonged devastating flooding in Sindh province. Mean monthly cases rose from approximately 10,900 (range 2,729–32,620) in the pre-flood period (January 2018– July 2022) to approximately 105,100 (range 31,759–250,746) from August 2022 onward. Since this shift reflected a discrete environmental and public-health disruption rather than continuous variation in the climate covariates of primary interest, it was modeled explicitly as a structural break using an interrupted time series (ITS) framework, rather than relying on the climate variables or a smooth time trend to absorb this discontinuity.

### Model specification

We fit a negative binomial generalized linear model (GLM) to monthly case counts. The negative binomial distribution was selected over a standard Poisson or quasi-Poisson specification because preliminary Poisson models exhibited substantial overdispersion. The negative binomial allows us to estimate a dispersion parameter (θ) directly rather than applying a post hoc variance correction to standard errors, making it a suitable model in this study.

The interrupted time series structure was represented by three terms: (1) a continuous, mean-centered time variable capturing the pre-existing linear trend; (2) a binary indicator for the post-flood period (August 2022 onward), representing an immediate level shift at the time of the break; and (3) a “time since flood” variable (coded 0 prior to the break, incrementing thereafter), allowing the post-break period to have its own linear trend distinct from the pre-break trend and from the level shift itself. This decomposition in the time series data follows standard segmented-regression practice for interrupted time series analysis, separating the “jump” from any subsequent change in slope [22]. This study design is usually used to study the effects of an intervention, disaster, or natural event on the occurrence of a disease over time [23].

We also incorporated seasonality in the model. Seasonality was represented using a first-order harmonic (paired sine and cosine terms with an annual period) rather than a full set of calendar-month indicator variables, in order to conserve degrees of freedom given the limited number of observations (n = 84 months; n = 82 after lagging) to maintain the accuracy, reliability, and unbiased nature of your parameter estimates

Climate covariates included monthly mean temperature (modeled with a linear and quadratic term to allow for a non-monotonic relationship, consistent with known non-linear effects of temperature on mosquito vector and parasite development) and monthly total rainfall (modeled linearly). One-month lagged terms for both temperature and rainfall were included to allow for a delayed effect consistent with the extrinsic incubation period of the malaria parasite and mosquito breeding-cycle dynamics.

### Addressing residual autocorrelation

An initial model specification (without an autoregressive term) showed significant first-order autocorrelation in the Pearson residuals (lag-1 autocorrelation ≈ 0.34, exceeding the 95% confidence bound), indicating that month-to-month dependence in case counts was not fully captured by the trend, seasonal, flood, and climate terms alone. To address this, the natural logarithm of the case count in the preceding month, log(cases[t-1] + 1), was added as a covariate. This log transformation was used because the model’s linear predictor operates on the log scale; including the lagged outcome in this form allows its coefficient to be interpreted as an elasticity (the proportional change in expected cases associated with a proportional change in the prior month’s cases).

Following inclusion of this autoregressive term, residual autocorrelation was no longer statistically significant at any lag, and model fit improved substantially (ΔAIC = 28.3; ΔBIC = 25.9, both favoring the autoregressive specification). This model was retained as the final specification.

### Final model

The final model can be summarized as:

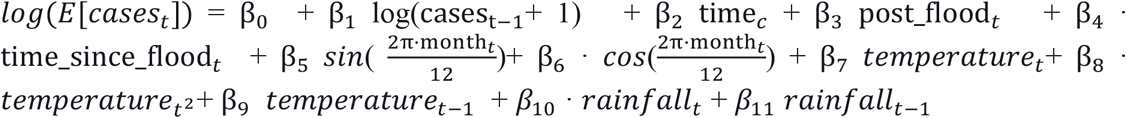

This model was fit via negative binomial regression (MASS::glm.nb in R), with *cases*_*t*_ ~ *NegBin*(*µ*_*t*_, *θ*).

### Testing for interactions

To assess whether the effects of climate covariates or seasonality differed before versus after the 2022 flood, or whether temperature and rainfall jointly modified each other’s effect, three candidate interaction terms were added individually to the final model and evaluated via likelihood ratio tests (LRTs) [24] against the model without interactions: (1) temperature × rainfall, testing whether the two exposures compound one another; (2) post-flood period × temperature, testing whether the temperature-case relationship differed after the flood; and (3) post-flood period × seasonal harmonic terms (sine and cosine components together), testing whether the shape of seasonality itself shifted post-flood.

None of the three interactions reached conventional statistical significance individually (temperature × rainfall: χ^2^(1) = 0.01, p = 0.92; post-flood × temperature: χ^2^(1) = 3.32, p = 0.068; post-flood × seasonality: χ^2^(2) = 5.40, p = 0.067), and none improved model fit by Akaike Information Criterion (AIC) by more than 1.4 points relative to the model without interactions. A joint model including all three interaction terms simultaneously reached nominal significance on a combined likelihood ratio test (χ^2^(4) = 10.90, p = 0.028) and showed a marginally lower AIC (Δ = 2.9), but had a higher Bayesian Information Criterion (BIC) than the model without interactions (Δ = 6.8), indicating that the added complexity was not justified once penalized for the number of parameters relative to the sample size (n = 83 months). Given the absence of a single robust, individually significant interaction and the BIC-based preference for the simpler specification, all interaction terms were excluded from the final model. This joint-significance-without-individual-significance pattern is interpreted as reflecting several weak, correlated effects rather than a single confirmed interaction, and is reported here as a formal robustness check with a null result rather than as a substantive finding.

### Model diagnostics

Model adequacy was assessed via: (1) comparison of Pearson residual variance between the pre- and post-flood periods, to confirm the negative binomial dispersion parameter adequately characterized variability in both regimes (observed variances of 0.95 and 1.11, respectively, indicating no meaningful discrepancy); (2) visual inspection of observed versus fitted values over time; (3) residuals-versus-fitted and residuals-versus-time plots to assess for remaining heteroscedasticity or trend; and (4) the autocorrelation function (ACF) of Pearson residuals, to confirm adequate resolution of temporal dependence following inclusion of the autoregressive term.

A non-parametric seasonal Mann–Kendall test was also performed to detect monthly trends of malaria, mean temperature and precipitation. This test is particularly useful to detect trends in data when underlying distribution is unknown. Mann–Kendall’s rank correlation tau and the associated p-value for temperature and precipitation was investigated [25]. The cross-correlation function between the time series of the weekly precipitation, weekly mean temperature, and the weekly malaria incidence assessed the time lags with peak correlations between malaria incidence and precipitation and mean temperature. These time lags were used in the final models of precipitation and mean temperature on the weekly malaria incidence. All analyses were performed in R (Version 2026.01.1+403 (2026.01.1+403)).

## Results

In this study, we explored the dynamics of the evolution of malaria incidence over time in relation to temperature and precipitation, on a pre- and post-flooding temporal scale. Fig. 1 presents the space-time evolution of malaria incidence over time in the study region. The results suggest that malaria risk increased substantially after the 2022 flooding in all the cities of the province. The final dataset for the analysis comprised 82 monthly observations (84 in total) from January 2018 through December 2024, following exclusion of two months with missing lagged covariates. Mean monthly malaria cases were 12641 (SD [15027]) before the August 2022 flood and 106481 (SD [63224]) after the devastating 2022 flooding. The majority of malaria-confirmed infections were Plasmodium Vivax (40-95%) between 2016-2024, followed by Plasmodium falciparum infections (Fig 3). The positivity rate for malaria remained <10% until June 2022, and increased massively after the July 2022 flooding (Fig 3).

**Fig 3:**
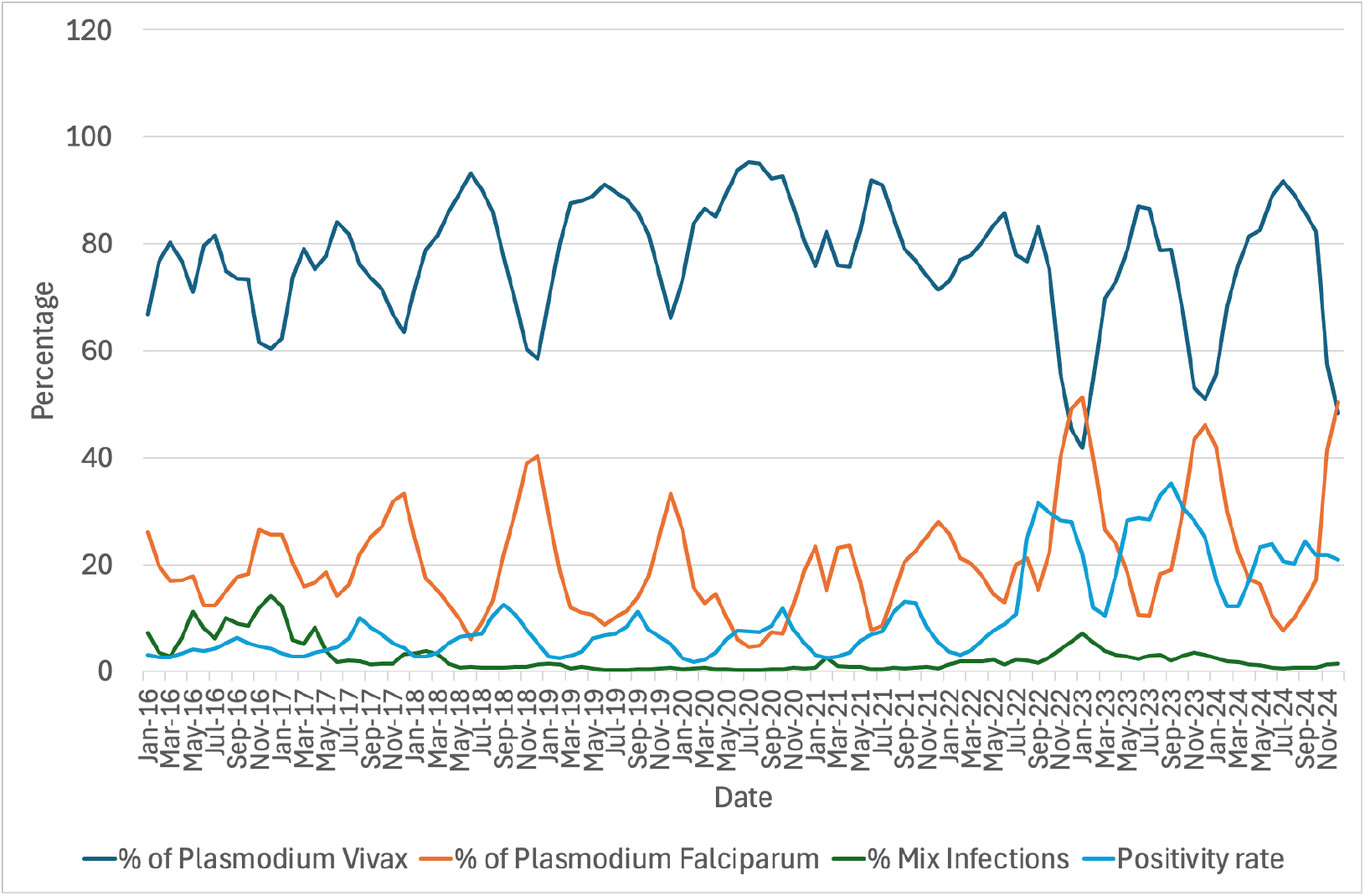
Percentage of Plasmodium Vivax, Plasmodium Falciparum, mixed infections (Plasmodium Vivax and Plasmodium Falciparum), and the malaria positivity rate in the population.

**Fig 4:**
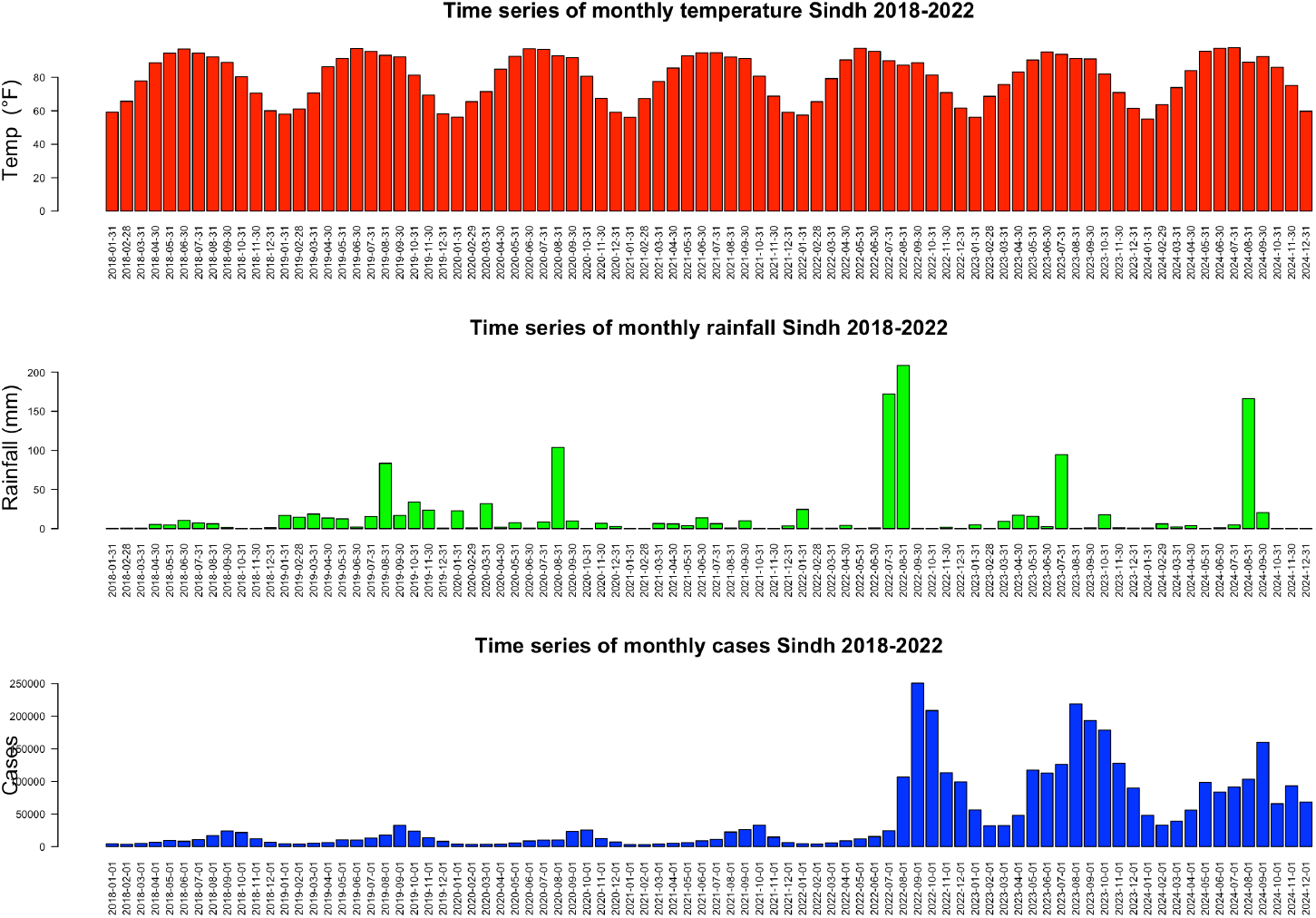
Panel plot showing the monthly mean temperature, monthly cumulative rainfall and monthly malaria case incidence in Sindh Pakistan from 2018-2024.

**Fig 5:**
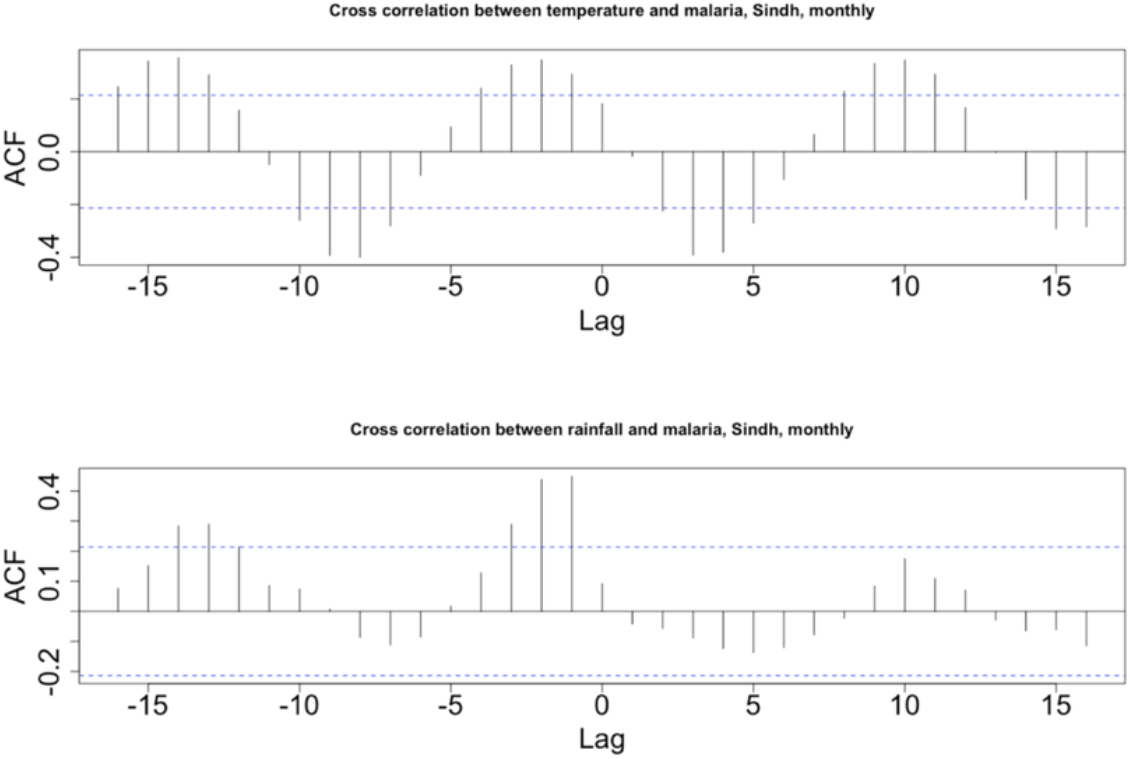
Cross correlations between temperture, rainfall and malaria cases.

The rainfall pattern in the province is highest between July and September, ranging up to 3 mm per month compared to 0 and 18 mm per month during the rest of the year. Sindh experiences a wet season from July to September and has a dry season for the rest of the year (~9 months). Average monthly temperatures in the province range from 55-97.5°F with the maximum temperatures reaching 111.6°F and the lowest hitting 43.3 °F.

The strongest positive cross-correlation between malaria cases and rainfall occurs at a lag period of −1 to −2 months (4-8 weeks), indicating that rainfall occurring 1-2 months prior to the current malaria cases resulted in the subsequent increase of malaria cases in the current month. The CCF of temperature and malaria showed scattered output. A negative lag of 1 month exists between malaria cases and temperature.

An interrupted time series model incorporating a first-order autoregressive term (the log of the prior month’s case count), harmonic seasonal terms, a pre-flood linear trend, a level shift at the onset of the 2022 floods, and climate covariates (temperature and rainfall, each with a one-month lag) provided a substantially better fit to the data than an equivalent model without the autoregressive term (AIC: [1646] vs. 1674 [no-AR1 model]; BIC: [1677] vs. 1703 [no-AR1 model]), both favoring the autoregressive specification). The negative binomial dispersion parameter (θ = 22.38, SE = 3.45) indicated the model adequately captured overdispersion in monthly case counts, and residual variance was similar in the pre- and post-flood periods (0.83 vs. 1.30; F-test for equality of variances, F=0.64, *p* = 0.16), indicating no evidence that the model fit differentially across the two periods. Fitted values closely tracked observed cases across the full study period, including the magnitude and timing of the post-2022 surge (Figure 1). An autocorrelation function of model residuals showed no remaining significant lag structure, confirming that the autoregressive term adequately addressed the temporal dependence present in an earlier model specification without it (Fig 6).

**Fig 6:**
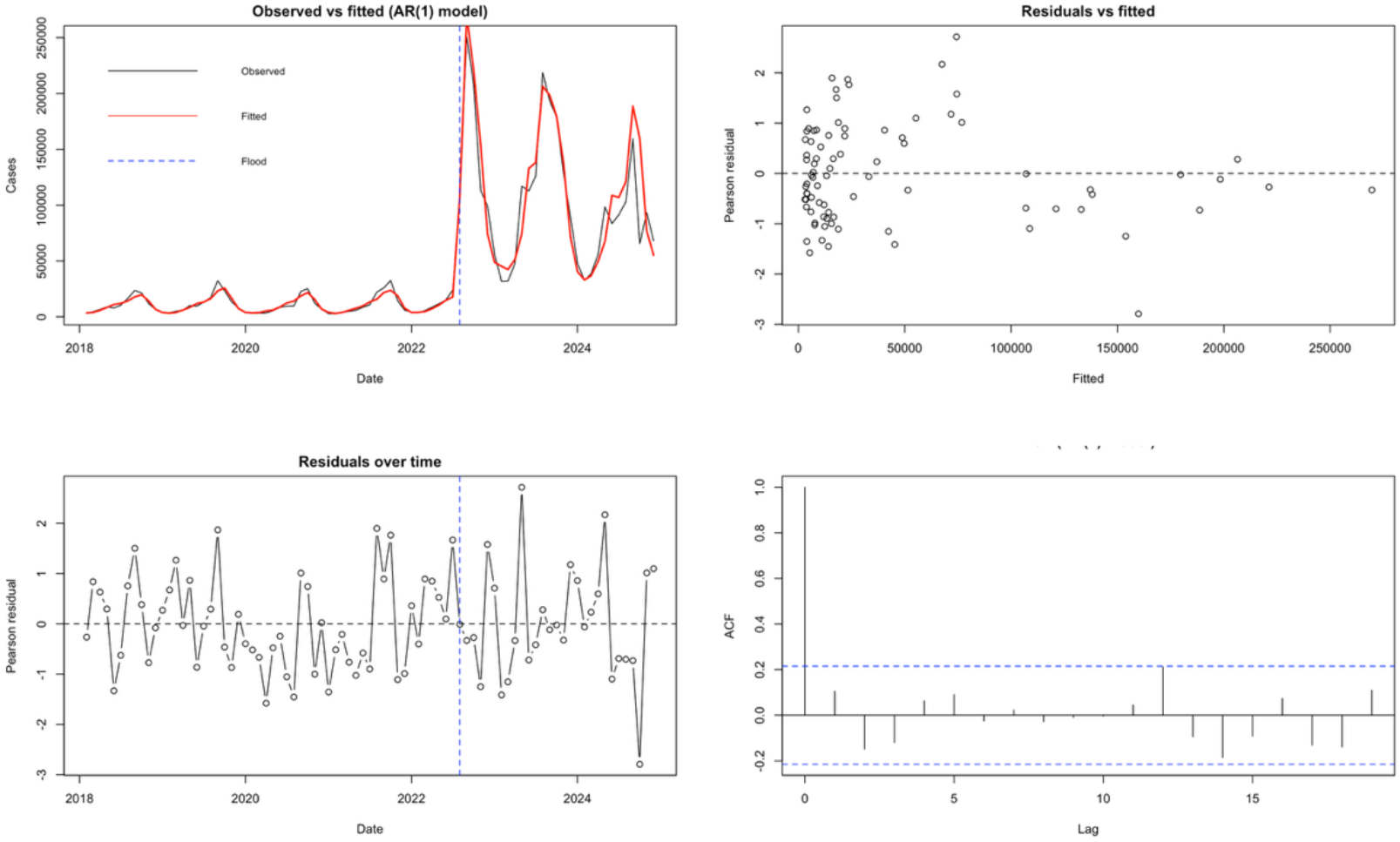
Panel plot showing the fit of the model, residuals and ACF over time.

The log of the previous month’s case count was strongly associated with the current month’s cases (incidence rate ratio [IRR] 1.696, 95% CI 1.431–2.008, p < 0.001). Because this term enters the model on the log scale, its coefficient can be interpreted as an elasticity: a 10% increase in the prior month’s cases was associated with an approximately 5.3% increase in the current month’s cases, holding trend, season, flood status, and climate covariates constant. This indicates substantial month-to-month persistence in transmission, consistent with ongoing local transmission chains rather than each month’s case count being driven independently by external conditions.

### Effect of Flood

After accounting for this case momentum, the onset of the 2022 floods was associated with an immediate, independent increase in malaria cases of approximately threefold (IRR 2.985, 95% CI 1.878–4.751, p < 0.001). This is substantially smaller than the roughly tenfold difference in raw means between the pre- and post-flood periods reported above, indicating that a sizeable portion of the apparent flood-associated increase reflects self-sustaining transmission momentum rather than a one-time exogenous shock alone. There was no evidence of an additional change in the underlying monthly trend following the flood beyond this level shift (time-since-flood term: IRR 0.990, 95% CI 0.979–1.001, p = 0.069), suggesting that the elevated post-flood case level has persisted rather than clearly resolving within the study period. The pre-flood period itself showed no significant secular trend (IRR 1.002 per month, 95% CI 0.999–1.006, p = 0.23). The post-flood trend showed a small, marginally significant decline of about 1% per month (IRR = 0.99, 95% CI: 0.979–1.001; p = 0.069), indicating the elevated post-flood level was largely sustained rather than rapidly decaying over the remaining study period.

For comparison, the model excluding the autoregressive term estimated a substantially larger immediate flood effect (IRR = 10.77 [CI not yet computed]). This divergence is consistent with the autoregressive term absorbing part of the flood’s effect through lagged case counts, since cases in the months following the flood were themselves elevated. Moreover, Log-transformed cases in the prior month were strongly predictive of current-month cases (IRR = 1.70, 95% CI: 1.43–2.01; p < 0.001), confirming meaningful month-to-month persistence in case counts beyond what seasonality and climate covariates alone explain (Fig 7).

**Fig 7:**
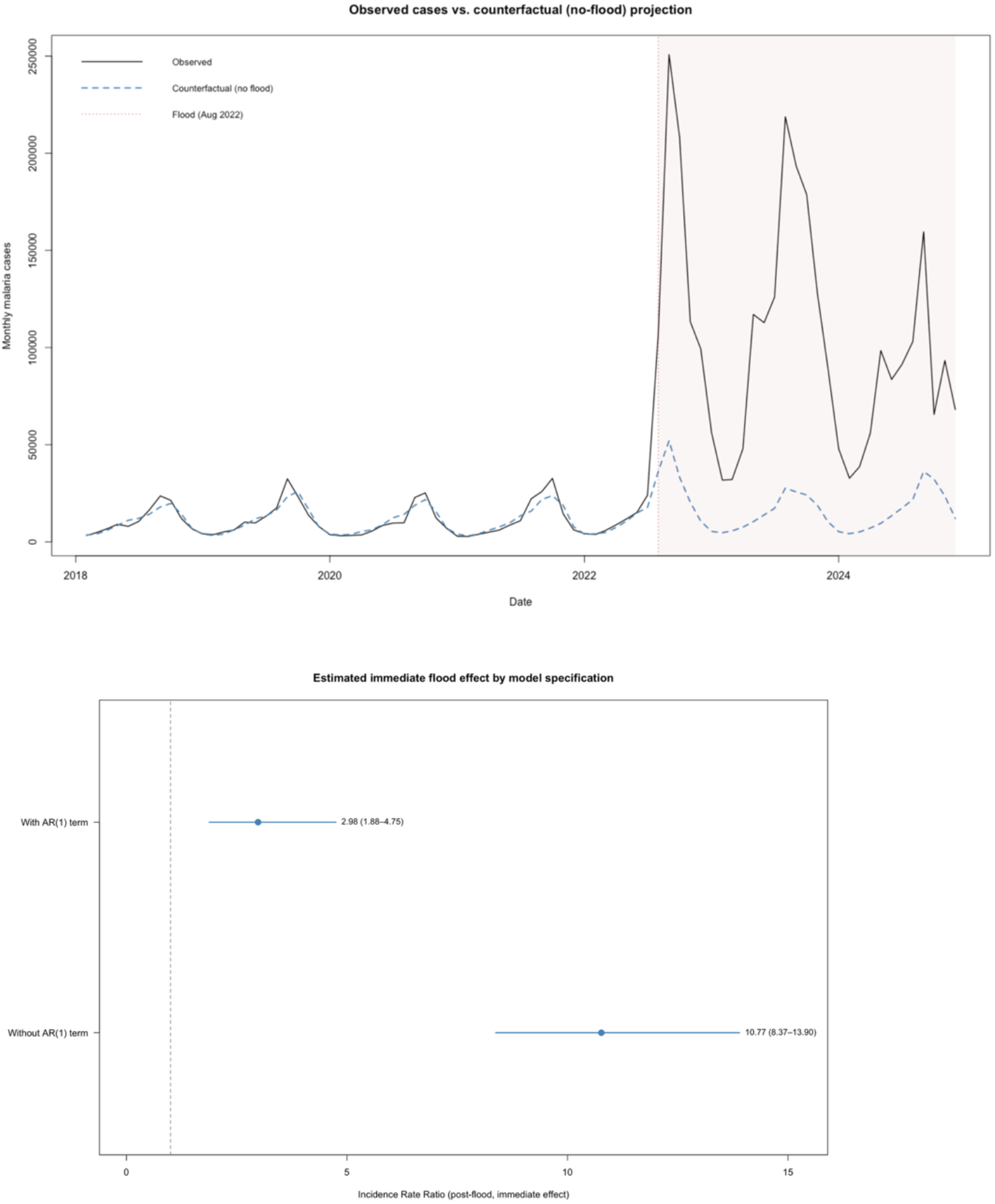
Impact of autoregressive lag term in the model.

### Impact of climate covariates on malaria cases

Mean monthly temperature was positively and significantly associated with malaria cases with a non-linear relationship. The linear temperature term was associated with increased incidence (IRR = 1.11, 95% CI: 1.04–1.17; p = 0.0014), while the negative quadratic term was negative and significant (IRR = 0.9994, 95% CI: 0.9989–0.9999; p = 0.012), indicating that incidence rose with temperature up to a point before plateauing or declining at the highest observed temperatures. The one-month lagged temperature term was not independently significant in this model (IRR 1.012, 95% CI 0.996–1.029, p = 0.15).

Monthly rainfall was not significantly associated with malaria incidence (IRR = 0.9996, 95% CI: 0.9979–1.0013; p= 0.63). Lagged rainfall (by one month) on the other hand was statistically significantly associated with malaria case incidence (IRR = 1.0030, 95% CI: 1.0012–1.0048; p <0.001), however, the magintude of the effect was small The underlying linear time trend (IRR = 1.002, 95% CI: 0.999–1.006; p = 0.23) was not statistically significant in the model.

### Impact of seasonality

Residual seasonality captured by the first-order harmonic term (sine and cosine) terms after accounting for trends, flooding and other climate covariates described above followed the expected annual cycle, with predicted relative risk lowest in the early months of the year and rising toward a peak in the mid-to-late months (Figure 8), consistent with the monsoon-associated transmission season in Sindh. Neither harmonic term was individually statistically significant at conventional thresholds (sine term: IRR 0.863, 95% CI 0.670–1.112, p = 0.27; cosine term: IRR 0.935, 95% CI 0.647–1.351, p = 0.73), which is expected given that sine and cosine terms jointly define a single seasonal curve rather than being independently interpretable. It suggests that the residual seasonal variation is largely captured by the lag-1 case term once it is included in the model. The Pearson residual variance is high post-flood vs. pre-flood (1.3 vs. 0.83), variance ratio 0.64 (*p*=0.161), indicating that after including the AR(1) term, there is no statistically significant remaining evidence of a pre/post-flood difference in residual dispersion—though the observed post-flood variance is still somewhat larger (Fig 8).

**Fig 8:**
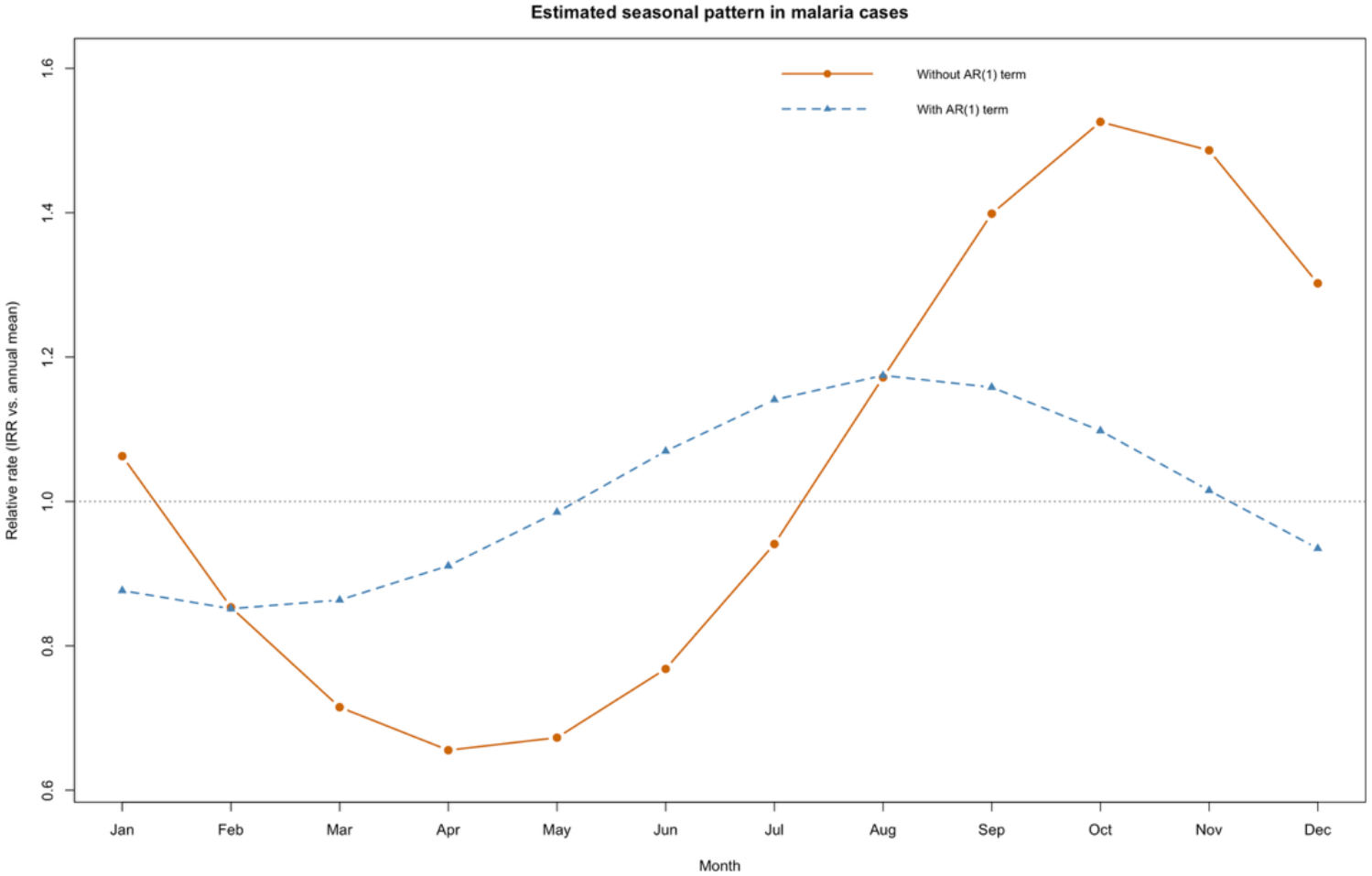
Estimated seasonal impact of rainfall and temperture on malaria cases in Sindh Pakistan.

### Interaction effects

Three pre-specified interaction terms; temperature * rainfall, flood period * temperature, and flood period * seasonality were tested individually via likelihood ratio tests against the base model. None reached conventional statistical significance individually (temperture*rainfall [p=0.9195], flood perid* temperature [p=0.06848], flood period * seasonlity [p=0.06721]) and a joint model including all three interactions was significant [p=0.02773]. This model was preferred by AIC but not by BIC relative to the base model without interactions (see Methods, *Testing for interactions*). No interaction terms were retained in the final model.

**Table:** AIC and BIC values of the model with and without the interaction terms.

| Interaction term | AIC | BIC |
|---|---|---|
| Model with no interactions | 1646.110 | **1677.555** |
| Temperature*rainfall | 1648.100 | 1681.963 |
| Flood period*temperature | 1644.719 | 1678.665 |
| Flood period*seasonality | 1644.710 | 1680.993 |
| Model with all interaction terms | **1643.212** | 1684.332 |

## Discussion

This study modelled the influence of selected climatic variables on the incidence of malaria in the province of Sindh, Pakistan over a period of seven years. We employed a negative binomial generalized linear model (GLM) and an interrupted time series approach to isolate the impact of flooding on malaria incidence in the region. Interrupted time series analysis has been utilized in malaria studies to quantify malaria diagnosis before and after the introduction of rapid diagnostic tests [26]. In our study, this method provides us an opportunity to account for the increase in malaria cases after the flooding event, as used in prior studies quantifying impacts of flooding on diseases such as Leishmania and diarrhea [23, 27].

The analysis has identified three principal drivers of malaria case incidence in Sindh Pakistan: a strong month-to-month case momentum consistent with ongoing local malaria transmission, a large and persistent increase in cases following the catastrophic 2022 monsoon floods as documented by scientists [19], and a non-linear temperature-incidence relationship consistent with the known thermal biology of Anopheles vectors and Plasmodium parasites [28]. In contrast, rainfall depicted at most a small and practically modest association with incidence once these other factors were accounted for. The results indicate a 3-fold increase in malaria cases (IRR 2.98, CI 1.88–4.75) after the flooding event in July 2022 that has not resolved within the subsequent two and a half years of available data. The 2022 monsoon floods were among the most severe in the country’s recorded history, submerging roughly a third of the country at their peak and disproportionately affecting Sindh province [29]. The public health mechanisms linking an event of massive flooding to a sustained malaria surge are well documented in the broader literature and plausibly include an expansion of mosquito breeding habitat from prolonged standing water, collapse of water, sanitation, and hygiene systems, large-scale population displacement into crowded, under-resourced settings, disruption of vector control and case-management infrastructure, and degraded surveillance and reporting capacity that may itself inflate recorded case counts independent of true incidence [29-31].

The application of an interrupted time series study design to this data, and specifically the inclusion of an autoregressive case-momentum term, allowed us to partially disentangle two related but distinct phenomena: an immediate, flood-associated increase in transmission risk (captured by the level-shift term), and the tendency of an established outbreak to sustain itself through ongoing local transmission independent of monthly climate conditions (captured by the autoregressive term). Flooding has been associated with increased risk of malaria in other countries including China, Mozambique, the Gambella region, and Sudan [32-36]. The finding that the flood-specific level shift (approximately threefold) was substantially smaller than the raw pre/post difference in means (approximately tenfold) indicates that a substantial share of the sustained elevation in cases reflects this self-sustaining transmission dynamic rather than a fixed, one-time environmental shock. This shows there is a high level of heterogeneity in the effects of weather on malaria transmission, as has been shown in prior studies [37]. This distinction has direct implications for intervention timing: measures aimed at breaking transmission chains (vector control, active case finding, prompt treatment) may have compounding value in an outbreak of this kind, beyond addressing the initiating environmental conditions alone.

The non-linear relationship identified between temperature and malaria incidence in this study corroborates with evidence from prior studies. In our study, malaria incidence rose with temperature before plateauing at the highest observed values. This is consistent with a substantial body of entomological and epidemiological literature describing an intermediate thermal optimum for malaria transmission [7]. This is reflective of the combined effects of temperature on mosquito survival [2, 38], biting rate, and the duration of the parasite’s extrinsic incubation period within the vector [38]. This pattern held even after controlling for the flood-related disruption and case momentum, strengthening the confidence that it is a genuine biological relationship rather than confounding by the concurrent trend.

The comparatively weak rainfall association is a notable point of divergence from some prior malaria-climate studies [39, 40], and from this study’s own preliminary models (see Methods), which found much larger and directionally unstable rainfall effects before the flood-period structural break and case-momentum term were incorporated. We interpret this as evidence that at least part of the apparent rainfall signal in unadjusted or under-specified models was confounded by the coincident timing of extreme rainfall and the flood-driven case surge, rather than reflecting a robust independent rainfall-transmission relationship at the monthly, province-wide scale examined here. This does not rule out a meaningful rainfall effect at finer spatial or temporal resolution (e.g., district- or week-level analyses), where the relationship between precipitation and vector breeding habitat may be more directly observable than in an aggregated monthly series spanning a large and hydrologically heterogeneous province.

These findings suggest that post-flooding malaria surveillance and response in flood-affected settings should anticipate not only an acute environmentally-driven surge in cases but also a subsequent period of elevated, self-sustaining transmission that may persist well beyond the resolution of the initiating environmental disruption, in this case the flooding. From a modeling perspective, this analysis illustrates the value of interrupted time series methods, and specifically of explicitly modeling case autocorrelation, when analyzing surveillance data spanning a major disruptive event, an approach that may be broadly applicable to other vector-borne disease series affected by extreme weather events, conflict, or other structural breaks. Future work should prioritize disaggregating this analysis to finer spatial resolution where flood exposure and rainfall are less confounded with province-wide reporting changes, validating the case definition and completeness of reporting across the pre- and post-flood periods, and extending the observation window as further post-flood data become available to better characterize whether and when the elevated transmission regime identified here begins to resolve.

## Data Availability

All data produced in the present study are available upon reasonable request to the authors

